# A portable molecular laboratory for rapid genotyping in the field: application to sickle cell disease

**DOI:** 10.64898/2026.05.05.26352080

**Authors:** Fabienne Grunder, Anne-Flore Hämmerli, Claude Isofa Nkanga Bokembya, Stéphen Hennart, Muriel Helmers, Naomi Azur Porret, Bertrand Graz, Cécile Choudja Ouabo, Hugues Abriel

## Abstract

**Background:** Sickle cell disease (SCD) is the most common recessive genetic disorder, caused by pathogenic variants of the *HBB* gene. SCD is associated with a range of clinical manifestations, including vaso-occlusive crises, infections, and severe anaemia, which contribute to increased morbidity and mortality. The frequency of pathogenic alleles is high in Sub-Saharan African countries, with heterozygous carriers reaching up to 25% of the population. Several methods can be employed for molecular diagnostics, with *HBB* gene sequencing being the most precise. However, access to DNA analyses and sequencing in Low- and Middle-Income Countries (LMICs), where SCD prevalence is high, is limited. Understanding genetic profiles is crucial at both individual and population levels, as it can guide public health strategies and facilitate accurate genetic counselling.

**Aim:** This feasibility study aimed to demonstrate that a portable medical genetic laboratory (in suitcases) can be used to genotype individuals for the *HBB* A, S, and C alleles and their combinations within a few hours outside of a laboratory setting.

**Methods and results:** We established a portable medical genetics laboratory capable of DNA extraction and isothermal DNA amplification using a commercially available kit for the A, S, and C alleles of the *HBB* gene. During one single study day, this portable lab was set up in a room where the Swiss Association of Patients with SCD was holding its annual meeting. We analysed the samples of 27 participants who were aware of their A, S, or C status. We collected buccal swabs and dried blood samples for genotyping. Genotype results for all participants were obtained within five hours after sample collection. In four cases, we observed discrepancies between the buccal swab and blood genotypes; three were resolved upon repeat testing, and one reflected donor chimerism following hematopoietic stem-cell transplantation.

**Conclusions:** This study demonstrates the feasibility and efficiency of using a portable medical genetics laboratory for rapid genotyping of *HBB* SCD alleles in community settings.This approach can improve access to molecular diagnostics in resource-limited environments. Such tools have the potential to significantly enhance local capabilities for genetic screening, counselling, and public health planning in regions heavily affected by SCD.

## Introduction

The burden of genetic diseases presents a significant yet often overlooked public health challenge, affecting a considerable portion of the global population. Among them, sickle cell disease (SCD) stands out as the most prevalent severe monogenic disorder in Africa and a significant cause of morbidity and early mortality. The latest Global Burden of Disease analysis estimated that approximately 7.7 million individuals were living with SCD worldwide in 2021, with the highest concentration of births and prevalent cases occurring in western and central sub-Saharan Africa, accounting for roughly six million of these cases (Esoh, Wonkam-Tingang et al. 2021, 2023). The disease is also prevalent in other regions, including the Americas and Asia (2023). Clinically, SCD is characterised by recurrent and excruciating vaso-occlusive episodes, coupled with chronic haemolytic anaemia, making it a substantial cause of early mortality. SCD is a recessively inherited monogenic disease caused by pathogenic alleles of the *HBB* gene, which encodes the beta-chain of haemoglobin (Inusa, Hsu et al. 2019).

Traditional medical genetic laboratories require sophisticated high-tech equipment, skilled personnel, and significant financial resources. Standard tools include next-generation sequencing platforms and polymerase chain reaction (PCR) platforms. These infrastructure demands have limited access for underserved populations; however, point-of-care and field-deployable technologies are expanding what is feasible outside conventional laboratories. Recent technological advancements have opened new possibilities for molecular testing beyond traditional laboratory environments (Nayak, Blumenfeld et al. 2017).

To our knowledge, no mobile applications have been developed specifically for medical genetics. In response, Xpedite Diagnostics GmbH, in collaboration with our research group, has created a customisable mobile laboratory for rapid genetic analyses. This portable unit features secure compartments for transporting instruments and reagents (Nayak, Blumenfeld et al. 2017), a stainless-steel workbench suitable for nucleic acid extraction and amplification, and a self-sufficient power system that enables independent operation. Complementing this, the company LaCAR (Liège, Belgium) has developed a CE-IVD certified kit for the rapid detection of *HBB* S and C variants, based on loop-mediated isothermal amplification (LAMP) technology (Detemmerman, Olivier et al. 2018). This method enables reliable genotyping of *HBB* sickle cell variants, thereby significantly reducing the time and complexity associated with traditional diagnostic approaches, such as haemoglobin electrophoresis.

The current study, PORTA-HBB, was designed as an initial proof-of-concept project to evaluate the feasibility of implementing this genotyping protocol in settings outside conventional laboratories. We conducted a one-day field trial with participants recruited from the “Association Suisse Drépano,” based primarily in the Romandie French-speaking region of Switzerland. Using the portable laboratory and LaCAR LAMP kits, we successfully identified *HBB* genotypes from both blood and buccal samples from all participants. These results demonstrate the feasibility, reliability, and user acceptability of deploying portable medical genetics laboratories for SCD diagnostics in real-world, non-clinical environments.

## Methods

### Study Design and Inclusion of Participants, Ethical Approval and Funding

A single-day feasibility trial was conducted in Lausanne with 28 participants recruited through the Association Suisse Drépano between March 10 and April 5 2025. Inclusion criteria included ages 16 to 80 (need of parent or guardian consent was waived by ethical committee) and prior knowledge of the *HBB* genotype. All participants signed a consent form. The study was approved by the Ethics Committee of Canton de Vaud (CER-VD) under project number <u>BASEC ID 2024-02503</u>. All procedures were conducted in accordance with the Declaration of Helsinki and the Swiss Human Research Ordinance (HRO). This research was funded through intramural grants from the University of Bern.

### Portable laboratory

The mobile laboratory was a self-contained, suitcase-sized unit designed for infrastructure-independent diagnostic workflows, particularly suited for point-of-care applications. This enabled immediate nucleic acid extraction and detection after sample collection. This portable solution is ideal for remote fieldwork, facilitating on-site diagnosis and timely intervention for diseases like neglected tropical diseases during screening campaigns, and for rapid outbreak management, as demonstrated by its successful use for Schistosoma diagnostics in Tanzania (Webster 2024). The portable laboratory offers several advantages, including full customisation, portability in a robust, air-transport compliant case, and independence from external facilities or power sources. Internal components included removable dividers, a stainless-steel workbench, a magnetic lid for organising consumables, a waste container, an LED headlamp/power bank, a thermobox for storage, magnetic tube racks, a lithium-ion battery with cables, and a solar panel for recharging. For this project, the suitcase was equipped with a MIC cycler (BioMolecular Systems, Upper Coomera, Australia) and a DC-AC converter, allowing it to operate on a car battery.

### Sample collection

Buccal samples were collected using ISOHELIX™ DNA/RNA Buccal Swabs (SK-1S). Participants were asked not to eat for one hour before sampling. Each swab was rubbed against the inner cheek mucosa for approximately 30 seconds on each side to collect epithelial cells. Samples were processed immediately after collection. Blood samples were collected by fingertip puncture using a lancet (Accu-Chek Safe-T-Pro Plus). Four drops of capillary blood were placed onto a Qiagen FTA Classic card. The blood spots were allowed to air dry at room temperature until further analysis.

### Sample Analyses

Genotyping for SCD was performed using the Hb S/C Kit (LaCAR MDx Technologies S.A, Belgium). This assay is based on the LAMP (Loop-mediated Isothermal Amplification) method, followed by thermal analysis and fluorometric detection (Detemmerman, Olivier et al. 2018). For dried blood spots, a 6 mm punch was transferred into 1 ml of lysis buffer. For buccal swabs, 1 ml of lysis buffer was added directly to the swab tube. Samples were vortexed briefly and incubated at room temperature for 10 minutes. Subsequently, 5 μl of the obtained lysate was added to PCR tubes containing 20 μl of reaction buffer. Positive (plasmid containing HbA, HbS and HbC alleles) and negative controls from the kit were also included. The LAMP reactions were carried out on a MIC Real-Time PCR instrument (BioMolecular Systems) under the following conditions: isothermal amplification at 68°C for 30 minutes, followed by melting-curve analysis from 35 to 80°C at a ramp rate of 0.2°C/s. Result interpretation was based on melting curve profiles generated by Mic. In the event of discrepancies between the participant’s announced status and our results, we repeated the analysis.

### Data Management and Analysis

Participant demographic and genotype data were recorded in REDCap (Harris, Taylor et al. 2009, Harris, Taylor et al. 2019). The analysis focused on genotyping success and the concordance between blood and buccal samples. Descriptive statistics and frequency distributions were applied. Study data were collected and managed using REDCap, an electronic data capture tool hosted at the University of Bern.

## Results

During a single study day (5 April 2025), the portable laboratory, consisting of two suitcases, was set up in a hotel meeting room where the Swiss Association of Patients with SCD was holding its annual event (Figure 1).

**Figure 1.**
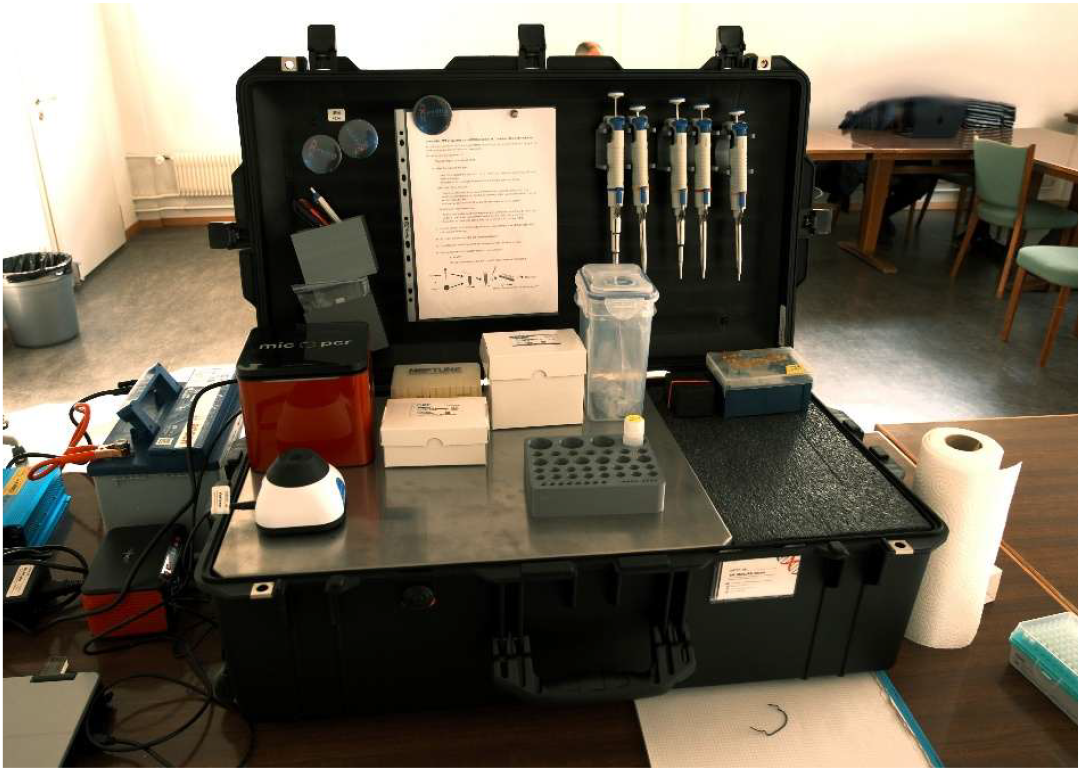
Picture showing the “portable laboratory” set up in the hotel meeting room, where sample collection took place and analyses were performed. On the left is the MIC PCR device used for the analyses.

We included 28 participants who were aware of their HbA, HbS, or HbC status. Two LAMP runs were performed to test buccal swabs and dried blood spots (DBS) from each participant, all of whom were informed of the study goals and provided written consent. Positive controls from the LaCAR kit showed the expected three melting peak temperatures for HbC, HbA, and HbS, whereas no peak was observed in the negative control (Figure 2). Allele-specific melting temperatures observed across samples were consistent with kit specifications (HbC ≈ 44 °C, HbA ≈ 51 °C, HbS ≈ 58 °C).

**Figure 2.**
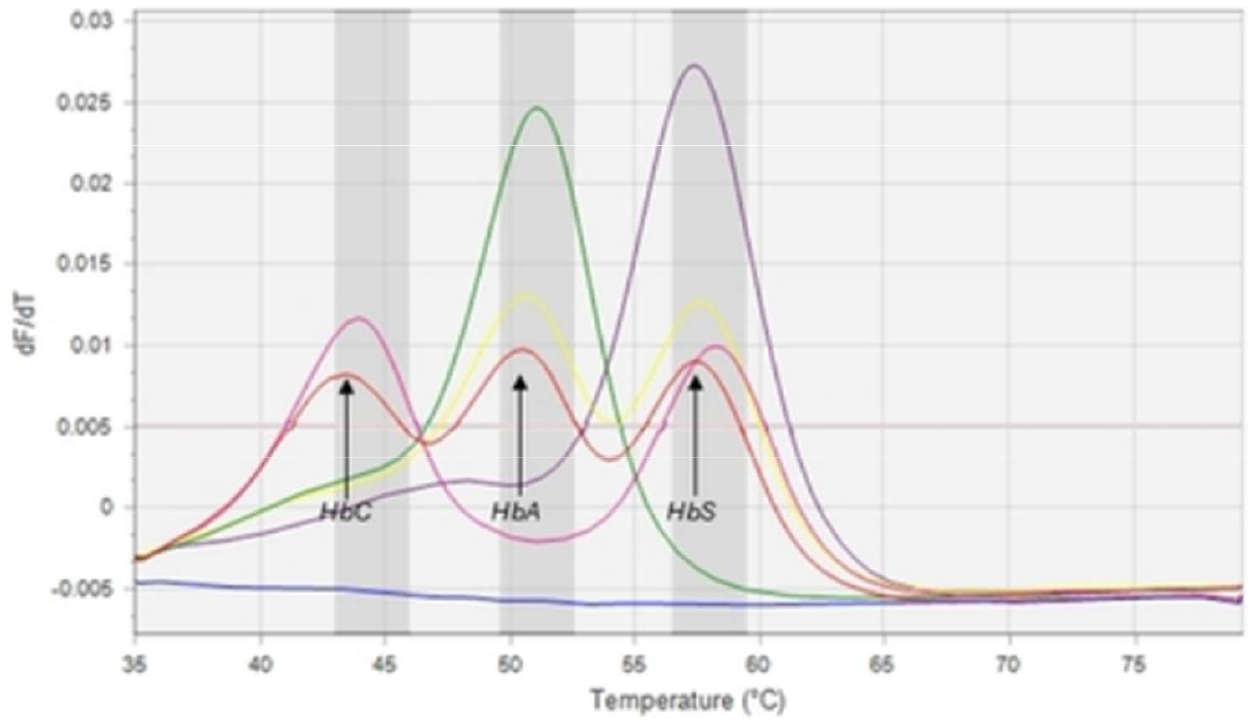
Representative melting-curve results from buccal swabs. Four participant genotypes are shown: HbA/A (green), HbA/S (yellow), HbS/S (purple) and HbS/C (pink). The negative control (blue) shows no peak, while the positive control (red) displays three allele-specific peaks corresponding to HbC, HbA, and HbS.

One participant was excluded *post hoc* because their *HBB* status was unclear. The genotypes identified among the 27 participants included HbA/A, HbA/S, HbS/S, and HbS/C. For the vast majority of participants, genotypes obtained from buccal swabs and DBS during the study day were concordant, except for 4 participants. Repeat testing 2-4 days after the study day resolved three of these discrepancies, most likely caused by pipetting errors. The remaining discrepancy was due to donor chimerism in a participant who had undergone hematopoietic stem-cell transplantation from an HbA/S donor, with buccal epithelial cells showing the recipient genotype (HbS/S) and blood cells reflecting the donor genotype (HbA/S). In total (Table 1), we identified 6 HbA/A, 14 HbA/S, 6 HbS/S, and 1 HbS/C among the DBS samples, and 1 less HbA/S and 1 more HbS/S among the buccal swab samples (see comment above regarding the grafted patient).

**Table 1.** Identified genotypes of the DBS and buccal swabs samples of the 27 participants (see text for more information).

| Genotype | Observed Genotype dried<br>blood spot samples | Observed Genotype<br>buccal swab samples |
| --- | --- | --- |
| AA | 6 | 6 |
| AS | 14 | 13 |
| SS | 6 | 7 |
| SC | 1 | 1 |

## Discussion

The main findings of this study were: (1) we demonstrated the feasibility of using a portable medical genetics laboratory for rapid SCD genotyping in a community setting. Genotyping for *HBB* alleles (HbA, HbS, and HbC) was completed in under five hours from sample collection to result interpretation, with high concordance between buccal swab and dried blood spot samples. (2) the demonstration that the LaCAR SCD genotyping kit can be applied to buccal swab samples without compromising accuracy, expanding its applicability to non-invasive sample types that are particularly advantageous in field settings and with paediatric populations.

In this feasibility study, we deployed the mobile laboratory at the annual meeting of the Association Suisse Drépano, providing a real-world environment to validate this decentralised testing approach. Among the 28 participants, one was excluded *post hoc*, and four initially showed discrepancies between the DBS and buccal swab samples. Repeat analyses resolved three of these discrepancies, indicating that they were most likely due to technical, handling, or sample-related issues during the initial runs. The remaining discrepancy involved a participant who had undergone hematopoietic stem-cell transplantation from a donor with an HbA/S genotype. In this case, buccal epithelial cells reflected the recipient genotype (HbS/S). In contrast, blood-derived cells showed donor chimerism (HbA/S), highlighting the assay’s ability to capture genuine biological differences between tissues rather than technical failure.

The LAMP-based assay proved to be a robust and rapid genotyping method. Compared with conventional PCR-based techniques, LAMP does not require thermal cycling, tolerates crude lysates, and is well-suited for portable platforms. Incorporating this chemistry into a fully self-contained laboratory—fitted in a suitcase and equipped with an autonomous power supply—presents an appealing model for molecular diagnostics outside of a laboratory setting. This study also confirms, for the first time, the use of non-invasive buccal sampling with the LaCAR kit without compromising the quality or reliability of the results. Buccal sampling provides a painless, quick, and infrastructure-light alternative suitable for field or outreach environments, such as schools, maternal clinics, refugee camps, and mobile health units.

Several limitations must be acknowledged. The study involved a modest sample size in a Swiss-based population already aware of their SCD profiles, which does not reflect real-world variability in low-resource settings. Larger validation studies in endemic regions with diverse populations are needed to confirm robustness, user training requirements, and long-term sustainability in routine care. Moreover, while the current assay targets the major *HBB* variants (HbS and HbC), future versions could include additional β-globin mutations, such as β-thalassemia and HbD and HbE variants, to address regional genetic diversity.

Nevertheless, the potential impact of this approach is considerable. SCD remains a significant cause of childhood morbidity and mortality in sub-Saharan Africa and other underserved regions. Early and accurate genotyping can enable timely clinical intervention (Houwing, de Pagter et al. 2019), reduce misdiagnoses, and improve reproductive counselling. This portable genetic diagnostic tool represents a scalable, context-adaptable solution for integrating genetic testing into national or regional screening programs and community-based health strategies.

In conclusion, this study confirms the feasibility of using a portable laboratory for field-based genotyping of SCD alleles, using both dried blood spots and, for the first time, buccal swabs. This innovation supports the broader goal of democratising access to genetic diagnostics, bridging infrastructure gaps, and advancing equitable genomic medicine worldwide. As precision public health advances, tools like this may play a key role in bridging diagnostic gaps and enabling equitable care for individuals with inherited disorders. Future research should focus on validation studies in endemic and resource-limited settings to further assess the analytical performance and reliability of this approach, including direct comparison with established gold-standard methods such as sequencing or conventional PCR-based assays.

## Data Availability

data are available upon request

## Acknowledgments

We are grateful to all the study participants (members and friends of the Suisse Drépano Association) for their voluntary involvement. We thank Josepha Lumpudi Nduhura Munga for her valuable assistance and presence on the study day. Special thanks to the technical and research staff at the University of Bern, Department of Clinical Research (CTU Bern), including Dr. Felix Rintelen and Dr. Alan Haynes, as well as to all the professionals and volunteers who contributed to the successful execution of the PORTA-HBB study day. The authors are grateful to Dr. Andy Wende for his collaboration in developing the portable laboratory prototype. HA is a member of the Genomics for Health in Africa Africa-Europe Cluster of Research Excellence, led by the African Research Universities Alliance (ARUA) and The Guild of European Research-intensive Universities.

## Notes

### Competing Interest Statement

The authors have declared no competing interest.

### Funding Statement

NONE

### Author Declarations

The study was approved by the Ethics Committee of Canton de Vaud (CER-VD) under project number BASEC ID 2024-02503.

